# Prevalence of Depression in pre- and post-operative CABG (Coronary Artery Bypass Graft) patients

**DOI:** 10.1101/2024.06.06.24308577

**Authors:** Maheen Tariq, Ismail Mazhar, Mir Muhammad Rai, Qudsia Umaira Khan, Muhammad Daniyal, Danyal Faisal

## Abstract

Depression in post-surgical patients is a common occurrence. It is likely to occur in cardiac surgery like Coronary Artery Bypass Grafting. This condition could significantly prolong recovery time and could result in complications. Therefore, this study aimed to study the prevalence of Post-Operative Depression amongst Coronary Artery Bypass Graft patients. The undertaken research was conducted amongst patients who had undergone CABG at CMH Lahore and Punjab Institute of Cardiology. A survey was conducted on a sample size of 150 patients, out of which 147 gave consent to participation. The sample size was calculated using WHO Formula. The severity of depression was analyzed through questions in the Becks Depression Inventory. Data was analyzed using SPSS software (25.0 version). Out of 147 patients, 110 were males and 37 were females. The results depicted greater depression amongst females than in males both pre-operatively and post-operatively, with a mean score of 1.41 and 1.08 respectively. A significant difference (p<0.01) was obtained between the correlation of Pre-Operative Scores (in percentages and frequency) and Post-Operative Scores (in percentages and frequency). An insignificant difference (p=0.125) was obtained between the correlation between Preoperative Scores and Gender. A greater Mean Score (1.41) was seen in females, indicating more post-operative depression than males (1.05). An insignificant difference (p=0.239) was obtained between the correlation between Preoperative Scores and Gender. A greater Mean Score (1.08) was seen in females, indicating more post-operative depression than males (0.84). The study concluded that the prevalence of postoperative depression among patients was not significant.

## Introduction

Depression is defined as an illness related to restlessness and anxiety, slowing down normal body functioning and thinking processes. It negatively affects a person’s physical and mental health. Depression is a major illness associated with many surgeries and the most common is Coronary artery bypass graft surgery (CABG). Coronary artery bypass graft surgery (CABG) is performed to treat coronary artery disease caused by the narrowing of the coronary arteries due to fat deposition, reducing the supply of oxygen-rich blood to the heart tissue. The risks that threaten patients undergoing surgery are postoperative bleeding, blood clot formation during the procedure, atherosclerosis, abnormal heart rhythms, and ultimately death. Post-operative depression leads to slow recovery of patients and also leads to more complications (1). WHO cites cardiovascular disease (CVD) and depression as the most two debilitating conditions in the health context, and these chronic diseases are among the diseases with the greatest impact on the quality of life (QOL) of any person (2).Depression is considered to be one of the main reasons for reduced well-being, harming a patient’s quality of life, as well as their social and family life. It is a strong risk factor for mortality as it is related to the occurrence of new cardiac events and reduced functionality up to six months post-CABG surgery, increasing the risk of hospital readmission in up to 20% of patients, due to complications including infection, arrhythmia, etc (3). This research aims to check the prevalence of depression and how it affects life activities in post-operative patients (4). Depression and coronary artery diseases are comorbid conditions with comorbidity approximately ranging from 14% to 47%. Many patients undergoing CABG, suffer from severe depression, both pre- and postoperatively (5). Age, gender, duration in Cardiac ICU, and the level of the hospital can affect the level of depression. Moreover, factors that increase anxiety during hospital stay after surgery including the hospital environment like taking antidepressant medications, concerns about personal things being inaccessible, and difficulty sleeping can be anticipated. Major depressive symptoms, although commonly encountered in medical populations, are frequently underdiagnosed and undertreated in patients with cardiovascular disease (6). Indicators that relate to postoperative depression, include cortisol, high-sensitivity C-reactive protein (hs-CRP), and oxidative stress biomarkers (7). Depression in post-operative CABG patients has several negative effects such as poor response to treatment, increased readmission frequency, slow recovery, and mortality (8). Recent studies reveal that adequate management of the psychological condition of Cardiac surgery and post-bypass patients will improve the quality of life and health outcomes in these patients (9). Depression after CABG decreases the activity of patients, leading to poor lifestyle and negative patient outcomes. Hence, it is necessary to diagnose and treat this condition, along with the cardiological disorders (10). Preoperative depression is predictive of decreased cardiac symptom relief, quicker return of symptoms, more frequent rehospitalizations, and increased mortality in the immediate postoperative period of depression (11). Anxiety and depression are common in patients undergoing coronary artery bypass graft surgery patients and are predictive of worse outcomes (12). This study aimed to correlate all factors to determine the prevalence of depression in CABG patients.

### Objectives

The main objective of this study was to check the prevalence of depression in post-operative Coronary Artery Bypass Grafting (CABG) patients. The target was to correlate major factors leading to depression in such patients.

## Methodology

The cross-sectional study was conducted in Punjab Institute of Cardiology, Lahore, Pakistan, and CMH Hospital, Lahore, Pakistan. Approval for conducting this study was received from the institutional review board of CMH Lahore Medical College, Pakistan. Adult patients who underwent Coronary artery bypass grafts from the mentioned hospitals in February 2023-October 2023 were included in the study. The exclusion criteria in this study were patients suffering from CABG complications. Patients who fulfilled the inclusion and exclusion criteria were enlisted in this study. The sample size of 150 participants was calculated using the WHO formula. Out of 150 patients, 147 gave consent for the study. Therefore, the cooperation rate was 98 %. No imputation method was used, and 147 patients were included for whom complete data was collected. All the participants were interviewed using the Beck depression inventory scale included in the survey. The interviews were conducted by two interviewers who were trained to ask questions to avoid interviewer bias. The interviewers were fluent in English and Urdu to avoid any communication errors. According to the score of depression, patients were placed into one of the seven categories of depression based on severity. The categories included No depression, these ups and downs are considered normal, mild mood disturbance, borderline clinical depression, moderate depression, and extreme depression. Each patient was interviewed before the operation to determine preoperative depression and was again interviewed one month after the operation to determine postoperative depression. The mean score of depression was used for analysis between preoperative and postoperative depression. Chi-Square test was performed to find differences in depression between preoperative and postoperative mean depression scores. Chi-Square was also used to find differences in depression scores based on gender. A p-value less than 0.05 was considered to show a significant difference.

## Results

A total of 150 patients were extended invitations to partake in the study, and 147 of them responded, resulting in a response rate of 98.00%.

## Figures

**Fig 1.**
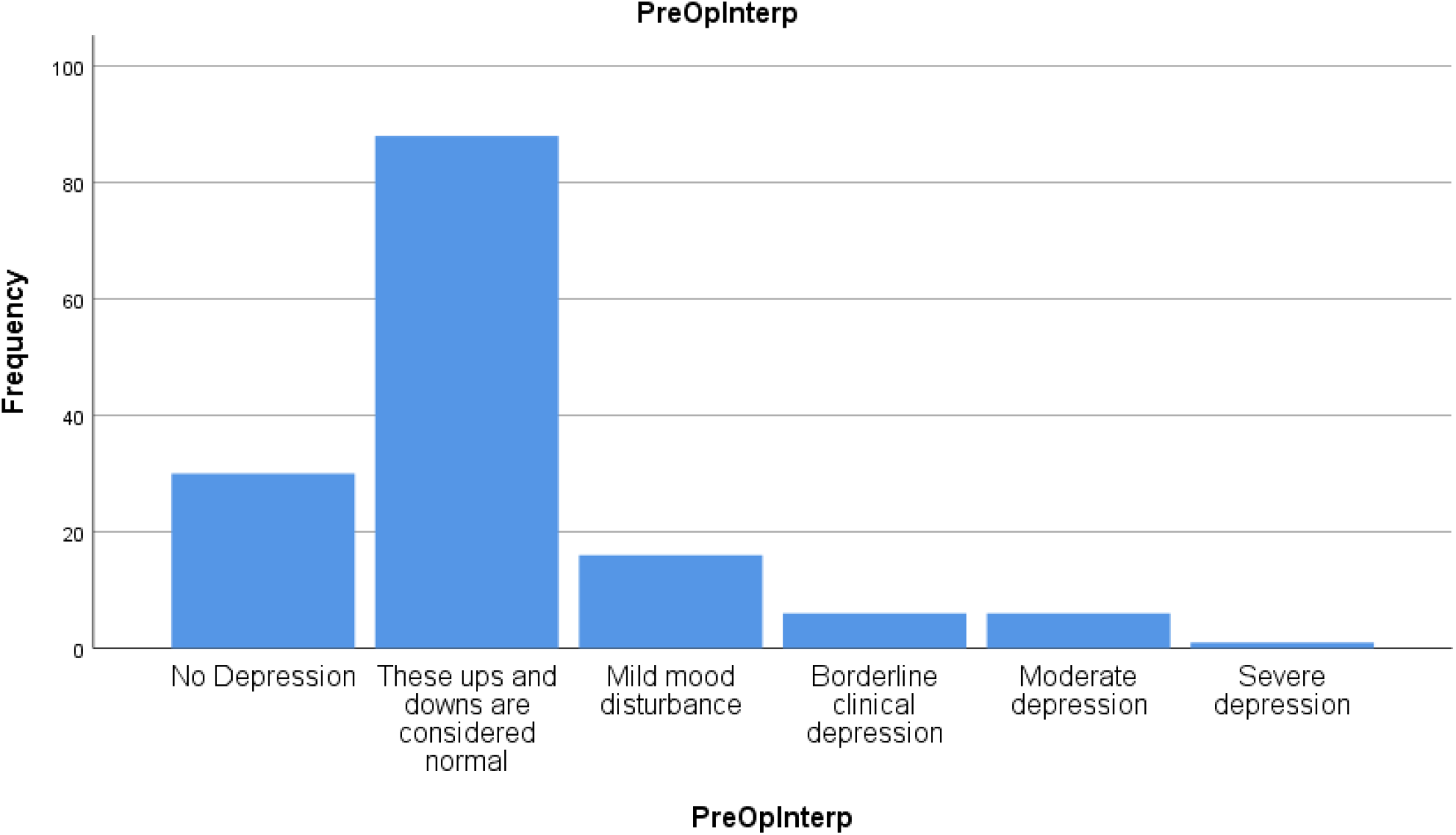
Pre-Op Descriptives.

**Fig 2.**
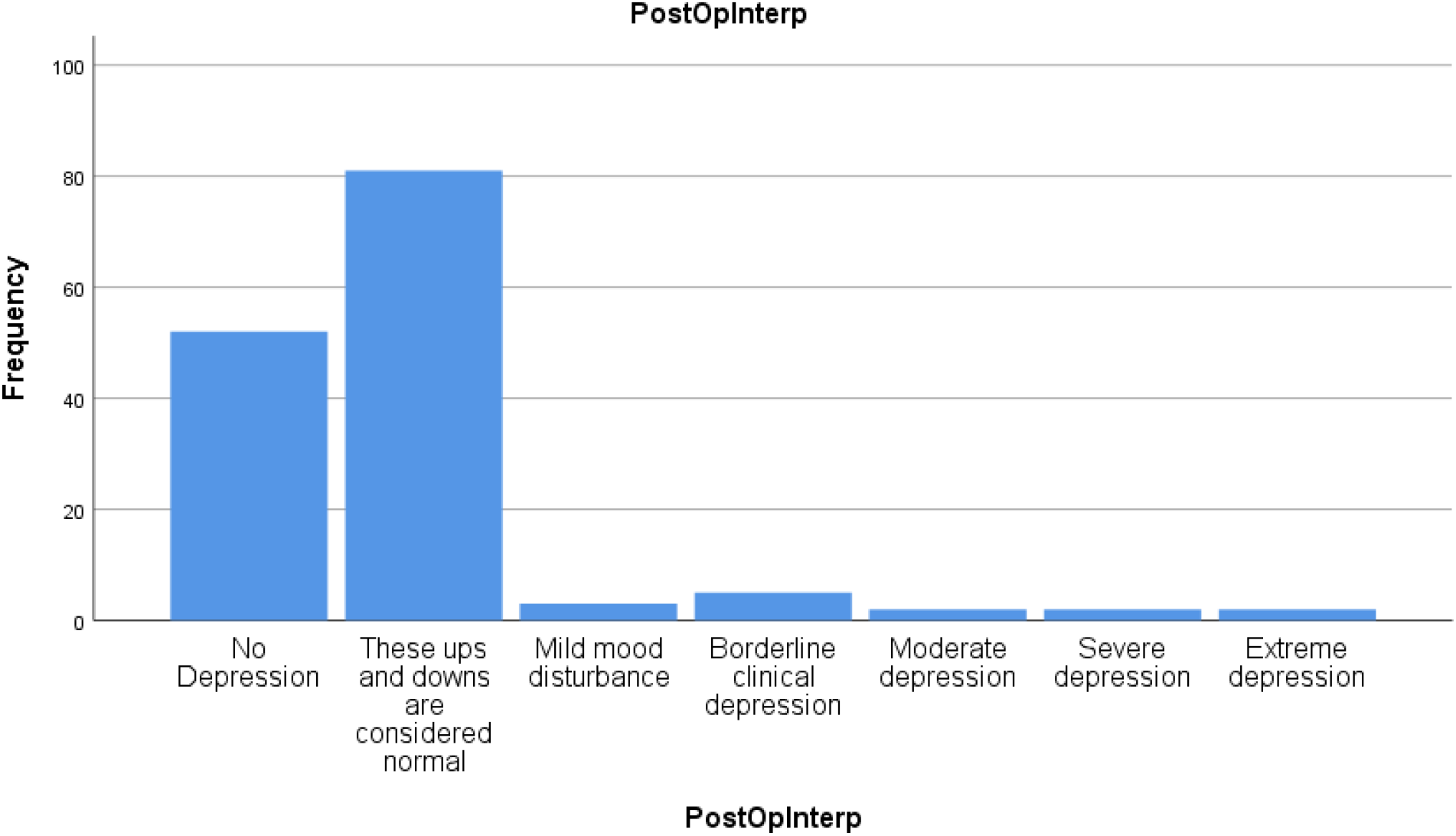
Post-Op Descriptives.

## Discussion

Our study aimed to check the Prevalence of Post-operative depression among patients undergoing CABG surgery. To achieve this, 147 people were asked to participate in the study. Table 1, classified these participants according to gender, with 110 males and 37 females.

**Table 1:** Demographic Variables.

| Gender | Frequency (n) | Percentage (%) |
| --- | --- | --- |
| Male | 110 | 74.8 |
| Female | 37 | 25.2 |

Among the participants, 110 (74.8%) were males, while 37 (25.2%) were females

Moving on to Table 2, the key finding of our study was that upon comparison of pre-operative and postoperative depression, there was a very significant difference, with a P-value of 0.001, in favor of depression being reduced in postoperative patients. This finding was further enhanced and lent support by a study in Amman, Jordan in which postoperative depression was also decreased in comparison to preoperative depression, with a similar p-value of 0.001 (13). As a result, these credible findings shed light on the statement that after the source of ailment was corrected or removed, quality of life improved and hence generated a more positive outlook on life whereas a lower quality of life and discomfort painted a more bleak picture.

**Table 2.** Descriptives and Correlation of Pre-Operative Scores with Postoperative Scores.

| Category | Pre-Operative Descriptives |  | Post-Operative Descriptives |  | Approximate Significance (P value) |
| --- | --- | --- | --- | --- | --- |
|  | Frequency | Percentage | Frequency | Percentage |  |
| No Depression | 30 | 20.4 | 52 | 35.4 | <.001 |
| These ups and downs are considered normal | 88 | 59.9 | 81 | 55.1 |  |
| Mild mood disturbance | 16 | 10.9 | 3 | 2.0 |  |
| Borderline clinical depression | 6 | 4.1 | 5 | 3.4 |  |
|  | Frequency | Percentage | Frequency | Percentage |  |
| Moderate depression | 6 | 4.1 | 2 | 1.4 |  |
| Severe depression | 1 | 0.7 | 2 | 1.4 |  |
| Extreme depression | 0 | 0 | 2 | 1.4 |  |
| Total | 147 | 100 | 147 | 100 |  |

A significant difference ( $p < 0.01$ ) was obtained between the correlation of Pre-Operative Scores (in percentages and frequency) and Post-Operative Scores (in percentages and frequency).

Similarly, in a meta-analysis regarding the topic of Depression after Heart Surgery, a significant p-value of 0.001 was once again observed regarding lowered depression post-operatively (14). Thus, further weight was lent to the veracity of our findings. On the other hand, depression could be a pre-existing condition as well in some cases, hence careful monitoring of the patient was required before and after the surgery. In other cases, it could be the hospital environment, and the behavior of the attendants, nurses, and doctors which could impact the prevalence of depression. Moreover, it emphasized the need for measures against depression and a proper intervention plan before and after the surgery (15).

Simultaneously, some studies also linked community and social support along with physical activity to reduced depression both preoperatively and postoperatively. One such study reported that in the Social Function (SF) domain, patients who were married, reported reduced mortality and shorter recovery times post-operatively along with optimism before the surgery. Along with that, in the Physical Function (PF) domain, patients who reported greater fears and depression before the surgery were mostly due to limitations in their daily activities, especially in those related to work (16).

In addition, even though it was not a parameter of our study, some articles also highlighted obesity as a factor that can impact the response before surgery. A South Asian study, which took responses mostly from obese patients reported a staggering prevalence of depression at 70.5%, along with lower physical activity also being an important cause, with a p-value of 0.05 (17).

Now coming to both Table 3 and Table 4, which placed gender as a defining parameter in regards to the prevalence, the snapshot portrayed that there was no significant difference in overall depression preoperatively, p-value of 0.125, and post-operatively, p-value of 0.239. However, in individual comparison of gender, a different picture was painted. Females displayed greater preoperative depression, with a greater mean score of 1.41, and postoperative depression, with a greater mean score of 1.08 in contrast to males. Furthermore, one other study also showcased greater depression in female patients preoperatively, but linked that depression to a longer hospital stay, with odds ratio being 4.04 times greater (18). In contrast, these findings were contradicted by another study in Kuala Lumpur where males were reported to have greater depression, with a p-value of 0.05 (19). This led to the perspective of whether biological differences alone were truly the differentiating factor concerning depression and there could be a multitude of other causes, especially those concerning quality of life and patient surroundings.

**Table 3.** Association of Pre-Operative Scores with Gender in CABG Patients.

| Gender | Frequency | Mean Score | Significance (P value) |
| --- | --- | --- | --- |
| Male | 110 | 1.05 | 0.125 |
| Female | 37 | 1.41 |  |

An insignificant difference ( $p = 0.125$ ) was obtained between the correlation between Pre-Operative Scores and Gender. A greater Mean Score (1.41) was seen in females, indicating more pre-operative depression than males (1.05).

**Table 4.**
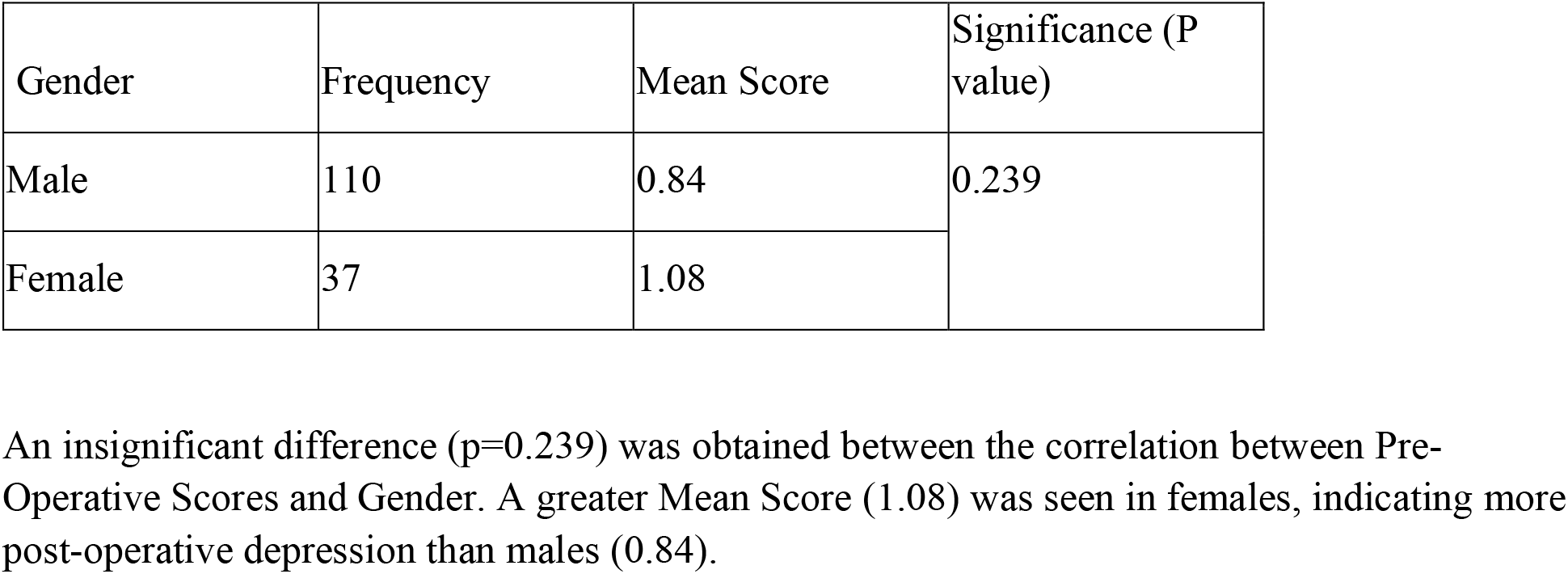
Association of Post-Operative Scores with Gender in CABG Patients.

| Gender | Frequency | Mean Score | Significance (P value) |
| --- | --- | --- | --- |
| Male | 110 | 0.84 | 0.239 |
| Female | 37 | 1.08 |  |

An insignificant difference ( $p = 0.239$ ) was obtained between the correlation between Pre-Operative Scores and Gender. A greater Mean Score (1.08) was seen in females, indicating more post-operative depression than males (0.84).

For future recommendations, proper psychological support and counseling must be provided to patients before the surgery and postoperatively to properly combat depression. Additionally, proper monitoring of patients regarding any risk factors. Moreover, it was significant to have proper rehabilitation programs in place to reduce recovery time and shorten hospital stays. Finally, hospital attendants must also be counseled to treat patients with more empathy and kindness.

Some of the limitations of our study were that:

- The samples obtained were restricted to a single hospital, hence decreasing the ability of our results to be generalized.
- The gender distribution displayed great disparity between males and females, hence decreasing equal representation of both categories in the result.
- Our study was limited to one parameter which was depression, and hence our result could not be further diversified.

## Conclusion

In conclusion, this study underscores the importance of recognizing and addressing depression in post-operative CABG patients, particularly among females, and advocates for integrated mental health care protocols as an integral component of cardiac surgery rehabilitation programs. The findings contribute to advancing our understanding of the psychological impact of CABG surgery and inform evidence-based strategies to enhance patient well-being and recovery outcomes.

## Data Availability

All relevant data are within the manuscript and its Supporting Information files.

